# Integrating Antimicrobial Stewardship and Infection Prevention Through Repeated Assessment and Feedback: A Multisite Quality Improvement Initiative in Viet Nam

**DOI:** 10.64898/2026.05.13.26353088

**Authors:** Quoc Phuong Nguyen, Van Giang Tran, Hai Yen Nguyen, Thi Phuong Oanh Pham, Trung Cap Nguyen, Minh Dien Vu, Anh Cuong Tran, Thi Ngoc Diep Nguyen, Van Minh Nguyen, Ba Hai Mai, Bao Dung Vo, Thanh Bao Nguyen, Dinh Phu Vu, Thi Thuy Van Pham, Thi Bich Ngoc Hoang, H Rogier van Doorn, Thomas Kesteman, Thi Lan Huong Vu

## Abstract

**Background:** Antimicrobial stewardship (AMS) and infection prevention and control (IPC) are complementary strategies to improve patient safety and address antimicrobial resistance (AMR). In low- and middle-income countries (LMICs), they are often implemented separately, reducing effectiveness. Evidence on integrating AMS and IPC in routine hospital practice remains limited.

**Objective:** To evaluate the feasibility of an integrated AMS-IPC improvement approach and describe changes in implementation in Vietnamese hospitals.

**Methods:** We conducted a multisite quality improvement initiative in four hospitals within the national AMR surveillance network in Viet Nam (March-September 2025). We used US-CDC’s tools to guide the implementation, including the Global Antibiotic Stewardship Evaluation Tool (G-ASET) and the Infection Control Assessment and Response (ICAR) tool. Baseline assessments were followed by feedback, multidisciplinary action planning, and targeted capacity building. Follow-up occurred 2-5 months later. Changes were analysed descriptively using quantitative scores and qualitative synthesis, and reported following the SQUIRE 2.0 guidelines.

**Results:** All hospitals had established IPC programmes at baseline, while AMS maturity varied. G-ASET scores improved across all sites, with greater gains in hospitals starting from lower baselines. Key improvements included leadership and governance, education and training, stewardship actions, and monitoring and reporting. IPC practices aligned with AMS priorities also improved, particularly transmission-based precautions, environmental cleaning, and cross-team coordination. Infrastructure-dependent areas, such as water safety, showed limited short-term progress.

**Conclusions:** An integrated AMS-IPC approach using repeated assessment and feedback is feasible and associated with meaningful improvements. This model offers a scalable strategy for strengthening hospital responses to AMR in LMICs and informs national programmes.

**Key messages:**

- Repeated assessment and structured feedback within an integrated AMS-IPC approach were associated with improved AMS implementation across diverse Vietnamese hospitals.
- Use of G-ASET and ICAR enabled hospitals to identify common system gaps and develop coordinated, locally tailored improvement plans.
- Gains were most evident in domains influenced by leadership, coordination, and behaviour change, whereas areas requiring financial investment showed less improvement over the short term.
- This assessment-guided network model may provide a scalable and cost-effective strategy to strengthen AMS-IPC integration in LMICs and inform future Fleming Fund-type investments.

**Summary of main point:** This study demonstrates that integrating antimicrobial stewardship and infection prevention through structured assessment and feedback is feasible in Vietnamese hospitals, leading to measurable improvements and offering a scalable, practical model for strengthening AMR responses in low- and middle-income countries.

## INTRODUCTION

### Problem description

Antimicrobial resistance (AMR) poses a major threat to patient safety and healthcare quality worldwide, with hospitals in low- and middle-income countries (LMICs) disproportionately affected. In hospital settings, inappropriate antimicrobial use and healthcare-associated infections (HAIs) interact to accelerate the emergence and spread of resistant organisms. Antimicrobial stewardship (AMS) and infection prevention and control (IPC) are core quality and safety strategies for addressing AMR and its consequences [1–3].

Despite this interdependence, AMS and IPC are frequently implemented as separate programmes, resulting in fragmented efforts, duplication of activities, and missed opportunities for system-level improvement [4]. This disconnect is particularly evident in LMICs, where IPC programmes are more widely established than AMS but are often under-resourced, and inconsistently implemented and limited by hospital infrastructure. Integration is further constrained by weak organisational structures, limited surveillance capacity, and workforce and diagnostic gaps [5]. Consequently, both IPC and AMS remain suboptimally implemented, with few integrated models in practice and limited evidence on their combined impact, underscoring the need for stronger, system-level integration [6,7].

In Viet Nam, national action plans on AMR and Ministry of Health guidelines mandate the establishment of multidisciplinary AMS committees and IPC programmes in hospitals [8–12]. However, implementation remains uneven. Studies from Viet Nam and other Southeast Asian countries have identified common barriers, including limited dedicated resources, insufficient training, weak governance structures, and suboptimal collaboration between clinical teams, pharmacy, microbiology, and IPC units [13–16]. As a result, AMS activities are often confined to guideline development or reporting requirements, while IPC efforts focus primarily on compliance with infection control standards, with limited coordination between the two.

Evidence further shows that although healthcare workers generally possess adequate IPC knowledge, adherence to recommended practices is variable, reflecting a well-described “know–do gap” [16,17]. Structural constraints, including infrastructure and design, overcrowding, limited microbiology capacity, and workforce shortages, further hinder effective implementation [17,18]. Assessments of IPC capacity across Vietnamese hospitals reveal heterogeneity in systems, lack of standardized procedures, and weak coordination between clinical and IPC teams [19]. These gaps contribute to ongoing transmission of HAIs and high levels of AMR, particularly in high-risk settings such as ICUs [20].

### Rationale

International guidance increasingly emphasises the integration of AMS and IPC as part of a systems-based quality improvement approach [21,22]. Integration has the potential to address shared drivers of AMR, such as leadership engagement, workforce capacity, surveillance, and management of priority AMR pathogens, more effectively than standalone interventions. However, evidence on how to operationalise AMS-IPC integration in routine hospital practice in LMICs is limited, particularly across hospitals with varying baseline capacity and resource constraints [15,23,24].

Viet Nam has developed a national AMR surveillance network, strengthened through international support, including from the Fleming Fund (2015-2025), which has enhanced laboratory capacity and expanded the availability of surveillance data [25,26]. Building on this foundation, there is a need to shift focus from data generation to implementation, supporting hospitals to translate surveillance and assessment findings into coordinated, locally relevant actions. Quality improvement approaches that combine repeated assessment, structured feedback, and context-adapted action planning offer a pragmatic pathway to achieving this integration, yet their feasibility and impact in real-world LMIC hospital settings remain underexplored.

### Aim

The aim of this multisite quality improvement initiative was to assess the feasibility of implementing an integrated AMS-IPC approach in routine hospital settings in Viet Nam, and to evaluate changes in AMS and IPC implementation following structured assessment, feedback and targeted capacity-building activities.

While the Fleming Fund has strengthened AMR surveillance capacity in Viet Nam, its conclusion without a subsequent phase [27,28] underscores the importance of sustainable, facility-level approaches, such as integrated AMS-IPC interventions, to maintain and operationalize these gains.

## METHODS

### Study design

This study reports a multisite quality improvement initiative integrating AMS and IPC through repeated assessment, structured feedback, and capacity-building interventions. The project was implemented within the Fleming Fund Phase II Country Grant in Viet Nam and followed principles of the SQUIRE 2.0 reporting guidelines for quality improvement studies [29] (Supplemental Data, Table S1).

The intervention used two complementary assessment tools:

- Global Antibiotic Stewardship Evaluation Tool (G-ASET): a structured, facility-level assessment instrument developed by the US CDC with the involvement of the G-ASET Development and Implementation Group in Asia [30–33] that evaluates the implementation of AMS programmes across core domains, using standardized indicators aligned with international AMS frameworks to identify gaps and guide targeted improvement.
- Infection Control Assessment and Response (ICAR): a practice-oriented and operational assessment framework developed by the US CDC to evaluate IPC capacity and practices across healthcare settings, covering key domains in alignment with WHO core components for IPC programmes [34,35].

Both tools were applied at baseline and follow-up to guide local action planning and evaluate changes over time.

### Setting and participating hospitals

The project was conducted between March and September 2025 in four hospitals within the Viet Nam Antimicrobial Resistance Surveillance Network, supported by the Fleming Fund. The hospitals were purposively selected to represent diverse healthcare contexts and baseline AMS-IPC capacity:

- National Hospital for Tropical Diseases (NHTD): a tertiary referral hospital specialising in infectious diseases.
- Viet Nam-Sweden Uong Bi Hospital (VSUH): a provincial general hospital in Quang Ninh province.
- Gia Lai Provincial General Hospital (GLPH): a provincial general hospital in the Central Highlands
- Hue University of Medicine and Pharmacy Hospital (UMPH): an academic general hospital in Central Viet Nam.

All four hospitals had existing IPC structures and microbiology services, but AMS programmes were at varying stages of development at baseline. Among these, two sites have previously been engaged in our AMS assessment research (2022-2023) at Oxford University Clinical Research Unit (OUCRU) - Viet Nam.

### Intervention overview: integrated AMS–IPC quality improvement approach

The intervention was designed as an integrated AMS-IPC quality improvement cycle, consisting of four core components:

1. Baseline assessment of AMS and IPC capacity using G-ASET and ICAR
2. Structured feedback to hospital leadership and multidisciplinary teams
3. Locally adapted action planning and capacity-building activities
4. Follow-up assessment to document changes and implementation progress

Integration between AMS and IPC was achieved through shared governance, coordinated action planning, and alignment of priorities, particularly around management of priority AMR pathogens (particularly those defined by WHO) [36], surveillance, and infection prevention practices. Overall timeline and activities are illustrated in Figure 1 and Table S2.

**Figure 1.**
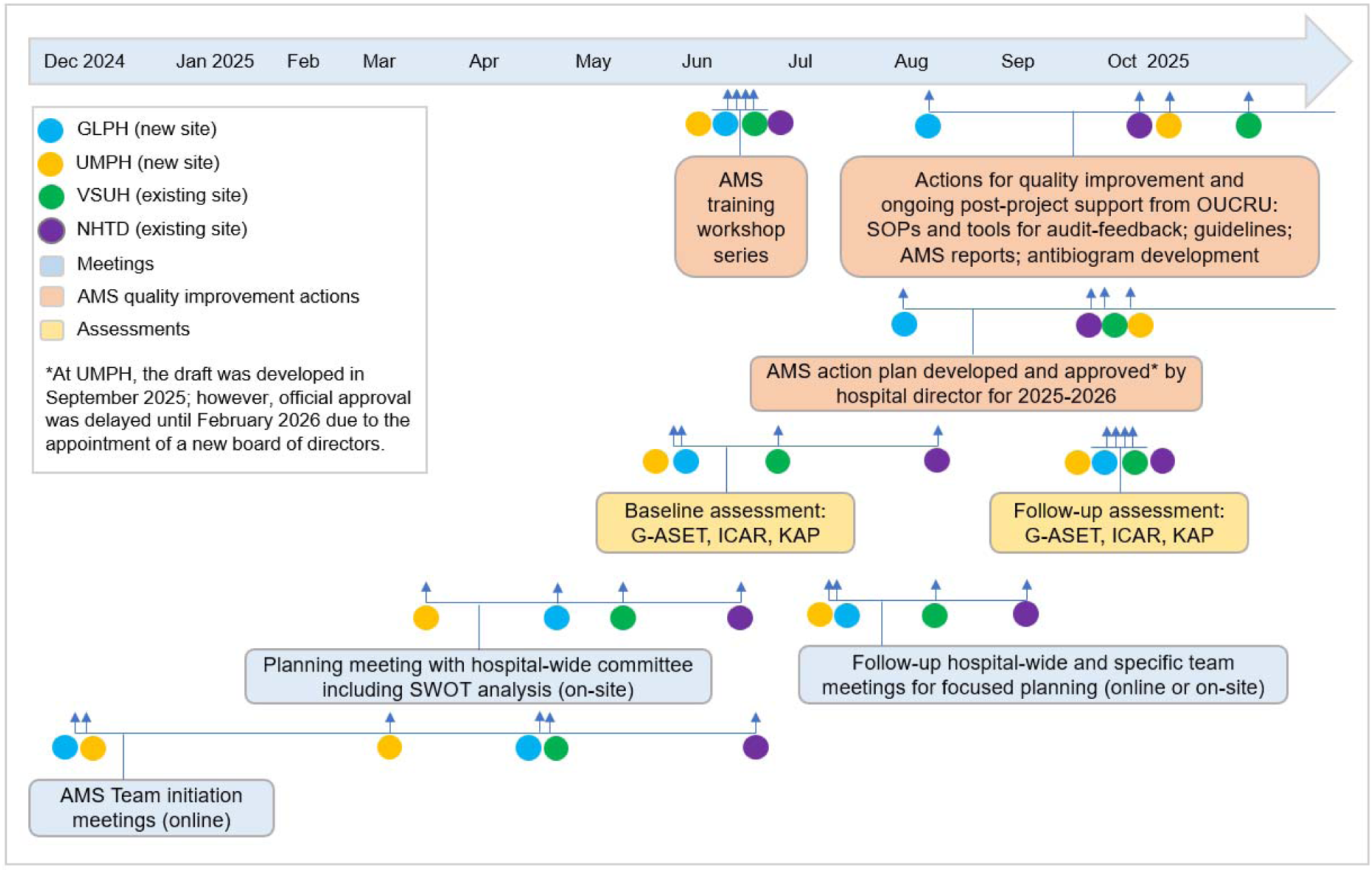
Timeline of integrated AMS-IPC activities across four hospitals, December 2024–October 2025. The figure shows AMS team initiation meetings, hospital-wide planning meetings with SWOT (strengths, weaknesses, opportunities, and threats) analysis, baseline assessments using G-ASET, ICAR, and KAP (knowledge, attitudes, and practices) tools, AMS training workshops, development and approval of site-specific AMS action plans, post-project quality-improvement support from the Oxford University Clinical Research Unit (OUCRU), and follow-up assessments using the same tools.

### AMS assessment: Global Antibiotic Stewardship Evaluation Tool (G-ASET)

AMS programme implementation was assessed using the G-ASET in five core domains of hospital AMS programmes:

- Leadership commitment and accountability
- Resources
- Education and training
- Specific AMS actions
- Tracking, monitoring, and reporting

The tool consists of 66 structured questions, with responses scored as: not implemented (0), partially implemented (2.5), or fully implemented (5).

Domain scores and overall scores were calculated for each hospital at baseline and follow-up. The G-ASET assessment was conducted through facilitated self-assessment meetings involving representatives from hospital leadership, clinical departments, pharmacy, microbiology, IPC, and quality management. To contextualise quantitative scores, semi-structured discussions with the research team were held during and after the assessment to explore barriers, facilitators, and contextual factors influencing AMS implementation. In addition, a knowledge-attitudes-practices (KAP) survey was administered by each hospital using the form developed under US CDC’s initiative for the G-ASET development [33] (Figure S1).

### IPC assessment: Infection Control Assessment and Response (ICAR)

IPC capacity and practices were assessed using the ICAR tool, focusing on the following domains under this quality improvement initiative:

- IPC programme structure and governance
- Hand hygiene
- Transmission-based precautions
- Environmental cleaning and disinfection
- Water safety and environmental sources of infection

ICAR assessments were conducted onsite by the hospital IPC teams, assisted by the study research assistant. The assessment combined document review, direct observation, and structured interviews with IPC personnel and clinical staff. Following each ICAR assessment, hospitals received written and verbal feedback highlighting strengths, gaps, and priority areas for improvement. Feedback was aligned with national Ministry of Health regulations and international guidance (US CDC and WHO).

### Action planning and capacity-building activities

Following baseline assessments, each hospital participated in multidisciplinary action-planning meetings involving hospital leadership, AMS committees, IPC teams, pharmacy, microbiology, and clinical departments.

Action plans were developed in alignment with:

- Viet Nam Ministry of Health (2020) Decision 5631/QĐ-BYT on hospital AMS implementation,
- WHO practical toolkit for AMS in LMICs (WHO 2019), and
- National IPC regulations (2017, 2018).

Capacity-building activities included the following main aspects (Supplemental Methods):

- targeted AMS training workshops (clinical pharmacy, microbiology, management of common infections),
- provision of AMS standard operating procedures and information materials,
- support for development or revision of hospital-level AMS-IPC action plans,
- tools to support prospective audit and feedback, including data collection templates.

AMS and IPC priorities were aligned, particularly for management of priority AMR pathogens, surveillance and reporting, isolation and transmission-based precautions, environmental and water safety measures influencing infection risk and antibiotic use.

### Follow-up assessment

Follow-up G-ASET and ICAR assessments were conducted approximately 2-5 months after baseline, depending on hospital scheduling and availability of key personnel (Figure 1). The follow-up assessments aimed to document changes in AMS and IPC implementation, assess progress against locally defined action plans, and identify persistent barriers and contextual constraints. Changes were evaluated descriptively by comparing baseline and follow-up findings within each hospital and across sites.

### Data analysis

Quantitative data from G-ASET were summarised using descriptive statistics, including domain-specific and overall scores at baseline and follow-up. Qualitative data from ICAR feedback, action-planning discussions, and assessment debriefs were analysed using thematic synthesis, focusing on: governance and leadership, workforce and training, integration between AMS and IPC activities, and contextual facilitators and barriers. Given the quality improvement design, no hypothesis testing or causal inference was undertaken.

### Ethical considerations

This project was conducted as a quality improvement initiative to support implementation of national AMS and IPC programmes. Formal ethical approval was obtained from the Ethics Review Committee at University of Oxford (OxTREC- 602-24) and NHTD Institutional Review Board (Decision 01-2025/HĐĐĐ-NĐTƯ). Participation was endorsed by hospital leadership, and no patient-level identifiable data were collected.

## RESULTS

### Baseline characteristics of participating hospitals

The four participating hospitals represented diverse healthcare contexts and baseline levels of AMS and IPC implementation (Table 1). National Hospital for Tropical Diseases, a tertiary infectious disease referral centre, demonstrated the most established AMS and IPC structures, including strong leadership engagement, multidisciplinary committees, and routine use of microbiology data to inform clinical and infection control decisions. The remaining three hospitals were provincial or academic general hospitals with existing AMS and IPC committees but more variable implementation capacity. At baseline, these hospitals had foundational governance structures in place but limited dedicated AMS resources, inconsistent formalisation of AMS activities, and uneven integration between AMS and IPC practices, particularly in education, surveillance, and coordinated management of priority AMR pathogens (Table 2).

**Table 1.** Baseline characteristics of participating hospitals. All hospitals had established AMS and IPC structures at baseline, although the level of operational maturity and prior exposure to formal AMS or IPC assessments varied across sites.

| Hospital | Level/ type | Geographic location | Primary clinical focus | AMS committee at baseline | IPC committee at baseline | Prior AMS/IPC assessment experience |
| --- | --- | --- | --- | --- | --- | --- |
| NHTD | Tertiary referral hospital | North Viet Nam | Infectious diseases | Established AMS programme with multidisciplinary committee and prior action plans | Established IPC programme with dedicated department and surveillance | Prior AMS assessments (US CDC initiative); no formal ICAR before this project |
| VSUH | Provincial general hospital | North Viet Nam | General acute care | Formal AMS programme in place but variable implementation in recent years | Established IPC programme with hospital-wide network | Prior AMS assessment (2023); no formal ICAR before this project |
| GLPH | Provincial general hospital | Central Highlands | General acute care | No formal AMS programme at baseline; antibiotic-related activities conducted without structured governance | Established IPC programme with routine practices | No prior formal AMS or ICAR assessment |
| UMPH | Academic general hospital | Central Viet Nam | General and teaching hospital | No formal AMS programme at baseline; partial stewardship activities embedded in clinical practice | Established IPC programme with variable operational maturity | No prior formal AMS or ICAR assessment |

**Table 2.** Baseline level of AMS and IPC programmes assessed using the Global Antibiotic Stewardship Evaluation Tool (G-ASET) and the Infection Control Assessment and Response (ICAR) tool. Baseline levels reflect overall patterns across hospitals rather than individual site scores.

| Domain | Assessment tool | Baseline level (overall) | Evidence from baseline assessments |
| --- | --- | --- | --- |
| Leadership and governance (AMS) | G-ASET | Moderate to high | AMS committees existed at all sites, but formal action plans and regular meetings were absent or inconsistent in three hospitals |
| Resources and workforce (AMS) | G-ASET | Moderate | Limited dedicated AMS staff and no protected budgets at most hospitals |
| Education and training (AMS) | G-ASET | Low to moderate | Lowest scoring AMS domain at baseline, particularly in provincial hospitals |
| Specific AMS actions | G-ASET | Moderate to high | Treatment guidelines and formulary controls present, but limited use of prospective audit with feedback to prescribers, and antibiograms in all hospitals |
| Tracking, monitoring, and reporting (AMS) | G-ASET | Moderate | Routine reporting existed, but limited feedback to prescribers and action planning |
| IPC programme structure | ICAR | Moderate to high | IPC departments and committees established at all hospitals |
| Hand hygiene | ICAR | High | Infrastructure and supplies generally adequate, but supply assurance, nail policies, hand-care support, and adherence monitoring varied across sites |
| Transmission-based precautions | ICAR | Moderate | Gaps in standardisation of signage, role clarity, and response to priority AMR pathogens |
| Environmental cleaning | ICAR | Moderate | Variable cleaning governance; two hospitals lacked formal product selection processes and two lacked minimum cleaning-time procedures |
| Water safety | ICAR | Low | Least mature IPC domain, with limited formal water safety programmes |

### Baseline AMS implementation assessed by G-ASET

At baseline, overall G-ASET scores demonstrated substantial variation in AMS programme maturity across the four hospitals. NHTD had the highest overall baseline score (84.9%), reflecting a well-established AMS programme with strong leadership engagement and routine stewardship activities. VSUH showed a moderate baseline score (78.0%), indicating the presence of a formal AMS structure but inconsistent implementation in recent years. GLPH and UMPH had lower baseline scores (both 73.5%), consistent with the absence of formally structured AMS programmes despite the presence of selected antibiotic-related activities. Across all hospitals, baseline scores were lowest in the domains of education and training and resources, highlighting common system-level gaps that informed subsequent capacity-building priorities.

Domain-level analysis of baseline G-ASET assessments revealed a consistent pattern of strengths and gaps across the four hospitals. Leadership commitment and accountability scored relatively high, reflecting the presence of AMS committees and senior management recognition of stewardship as a priority, although regular committee meetings and formally endorsed action plans were absent or inconsistently implemented in three hospitals. Specific AMS actions, including the existence of treatment guidelines, formularies, and selected stewardship activities, were also comparatively well developed at baseline. In contrast, education and training represented the weakest domain across sites, particularly in provincial hospitals, with limited structured AMS training for clinical staff, students, and trainees. Resource-related gaps were also evident, including a lack of dedicated AMS personnel, protected time, and formal budget allocation. Tracking, monitoring, and reporting activities were present in all hospitals but were often focused on routine reporting requirements, with limited use of audit findings, feedback to prescribers, or systematic action planning to optimise antibiotic use.

### Baseline IPC implementation assessed by ICAR

Baseline ICAR assessments indicated that all four hospitals had established IPC programmes, including formal committees or networks, designated staff, written IPC procedures, access to electronic medical records, and routine IPC risk-assessment activities. Hand hygiene was the most consistently established domain: all hospitals reported alcohol-based hand rub use in most clinical situations and soap-and-water handwashing for visibly soiled hands. Standard healthcare-worker hand hygiene indications were reported across sites, although supply assurance mechanisms, nail-related policies, staff hand-care support, and monitoring of adherence varied.

Transmission-based precautions, particularly for priority AMR pathogens, were reported across hospitals but varied in implementation, including differences in signage, role clarity, and isolation capacity. One hospital reported reliance on cohorting and care sequencing when single rooms were unavailable. Environmental cleaning systems were present, but governance was uneven; two hospitals lacked formal cleaning-product selection processes and two lacked minimum cleaning-time procedures. Water safety was the least mature IPC domain: only one hospital had a clearly dated water-related IPC risk assessment, three lacked sink-drain biofilm-reduction measures, and none had procedures for planned water-supply interruption. These baseline IPC gaps, particularly in transmission control and environmental domains, were identified as priorities for alignment with AMS interventions during subsequent integrated action planning.

Taken together, baseline G-ASET and ICAR assessments showed that while foundational governance structures for both AMS and IPC were present across all hospitals, important operational gaps persisted, particularly in education, workforce capacity, feedback mechanisms, and standardisation of transmission-based and environmental controls. These convergent weaknesses highlighted the interdependence of AMS and IPC and provided a clear rationale for an integrated quality improvement approach that aligned stewardship activities with infection prevention priorities.

### Integrated AMS–IPC action planning and implementation

Following baseline assessments, all four hospitals undertook multidisciplinary action-planning processes that explicitly integrated AMS and IPC priorities. Action-planning meetings involved hospital leadership, AMS committees, IPC teams, pharmacy, microbiology, and clinical departments, and were guided by findings from both G-ASET and ICAR assessments. These discussions enabled hospitals to prioritise system-level gaps that affected both antibiotic use and infection transmission, particularly governance, workforce capacity, education, surveillance, and management of priority AMR pathogens.

All hospitals developed context-specific AMS action plans that incorporated IPC-related activities, including strengthened collaboration between AMS and IPC teams, assignment of IPC representatives to AMS committees, and alignment of stewardship activities with transmission-based precautions for priority AMR pathogens and environmental hygiene practices. Three hospitals (NHTD, VSUH, and GLPH) obtained formal approval of their AMS action plans from hospital leadership during the project period, while the action plan at the remaining hospital was finalised but its approval was delayed by five months because of ongoing administrative transitions. Across sites, action plans commonly prioritised establishment or strengthening of core AMS teams, regular committee meetings, structured education and training, review and updating of treatment guidelines, prospective audit and feedback, improved use of microbiology data and antibiograms, and enhanced coordination with IPC activities such as hand hygiene monitoring, cohorting and isolation practices, and surveillance of hospital-acquired infections.

Capacity-building activities supported implementation of these integrated plans and included targeted AMS training workshops, dissemination of standard operating procedures, and provision of tools to support prospective audit and feedback and routine monitoring. Integration between AMS and IPC was operationalised through shared governance mechanisms, joint review of surveillance data, and coordinated feedback to clinical teams, particularly for infections caused by priority AMR pathogens. These processes formed part of a structured quality improvement approach, laying the foundation for improvements observed at follow-up and enabling hospitals to tailor AMS and IPC activities to their local contexts. The intervention was delivered through a multi-hospital network, enabling shared learning, peer comparison, and coordinated capacity building across sites.

### Changes in AMS and IPC implementation at follow-up

Follow-up assessments conducted 2–5 months after baseline demonstrated improvements in AMS implementation across all four hospitals, with the magnitude of change varying by baseline maturity and local context. Overall AMS implementation scores increased at all four hospitals between baseline and follow-up, with the largest improvements observed at sites with lower baseline maturity (Table 3).

**Table 3.** Overall AMS programme implementation scores at baseline and follow-up across four hospitals, assessed using G-ASET. All hospitals demonstrated increases in overall scores, with larger improvements observed at sites with lower baseline maturity.

| Hospital | Baseline<br>G-ASET overall<br>score (%) | Follow-up<br>G-ASET overall<br>score (%) | Absolute change<br>(percentage<br>points) | Time between<br>assessments |
| --- | --- | --- | --- | --- |
| NHTD | 84.9 | 86.4 | +1.5 | 2 months |
| VSUH | 78.0 | 95.5 | +17.5 | 3 months |
| GLPH | 73.5 | 90.1 | +16.6 | 5 months |
| UMPH | 73.5 | 88.6 | +15.1 | 5 months |

Domain-specific analysis (Table 4) indicated consistent improvements in leadership and governance, education and training, and tracking and reporting. Hospitals strengthened AMS committee functioning through more regular meetings, clearer role assignment, and formalisation of annual action plans. Education and training scores improved following targeted AMS workshops, particularly in hospitals with limited structured training at baseline. Improvements were also observed in tracking, monitoring, and reporting, including greater use of prescription audits, development or updating of antibiograms, and increased feedback of stewardship findings to clinical teams. Persistent challenges remained in securing dedicated financial resources and protected staff time for AMS activities, particularly in provincial hospitals.

**Table 4.**
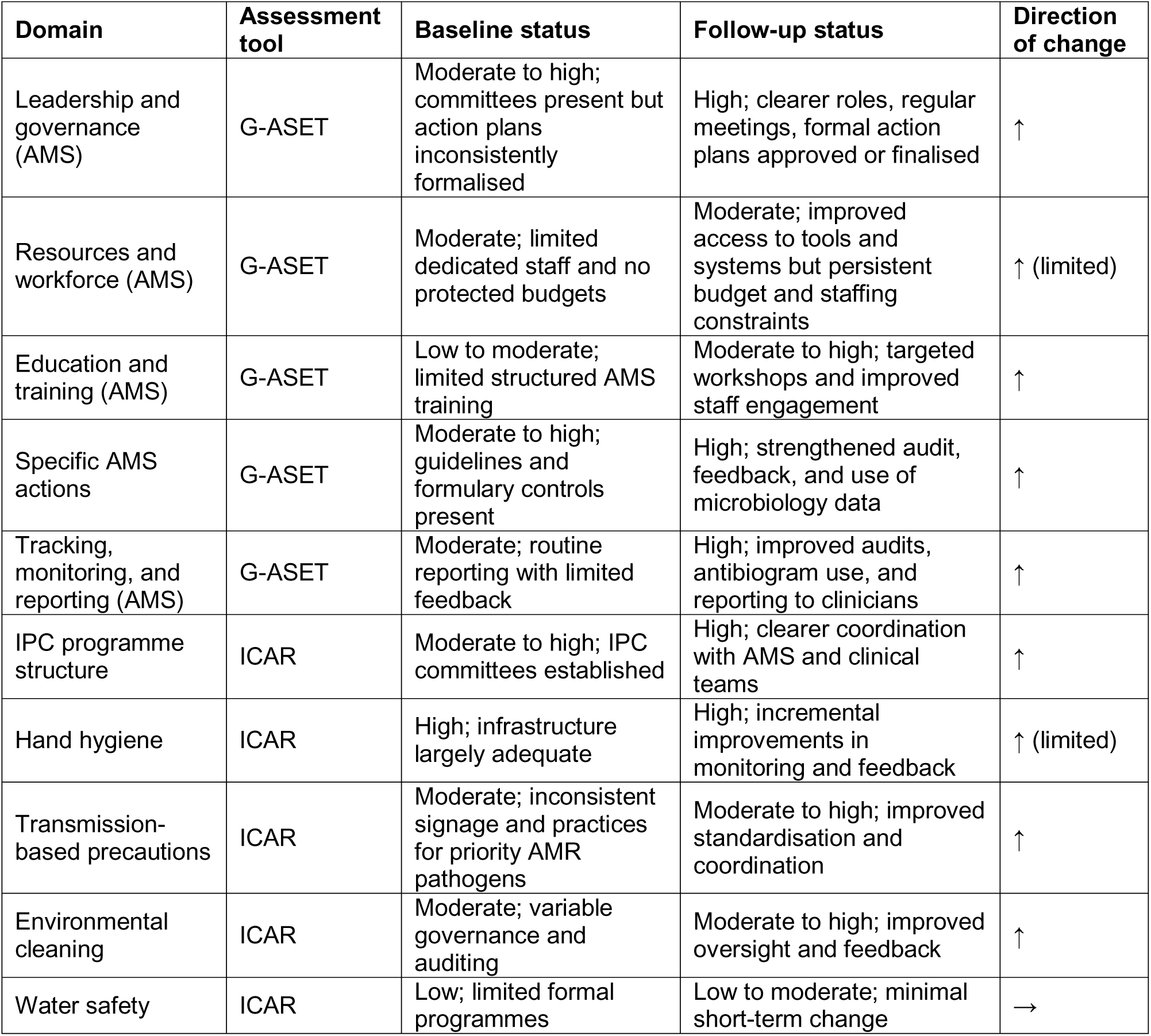
Baseline and follow-up changes in AMS and IPC domains, assessed using G-ASET and ICAR.

Parallel improvements were observed in IPC practices at follow-up, particularly in areas aligned with AMS priorities. Hospitals reported clearer implementation of transmission-based precautions, improved coordination between IPC, microbiology, and clinical teams for management of priority AMR pathogens, and more systematic feedback on hand hygiene and environmental practices. While foundational IPC structures remained stable, incremental gains were noted in standardisation of isolation practices and environmental cleaning oversight. Water safety practices showed limited short-term change, reflecting structural and infrastructural constraints beyond the project timeframe.

Overall, follow-up findings indicate that integrated action planning and capacity-building activities were associated with measurable improvements in AMS implementation and selected IPC practices, particularly those requiring governance, coordination, and behavioural change rather than major infrastructure investment.

## DISCUSSION

This multisite quality improvement initiative shows that an integrated approach to AMS and IPC [21,22], supported by repeated assessment, structured feedback, and targeted capacity building, was associated with measurable improvements in hospital AMS implementation across diverse healthcare settings in Viet Nam. By applying complementary assessment tools (G-ASET and ICAR), hospitals were able to identify convergent gaps in governance, workforce capacity, education, and feedback mechanisms, and translate these into locally adapted action plans that aligned stewardship activities with infection prevention priorities. Improvements were greatest in hospitals with lower baseline status of AMS implementation and more modest in an already well-established tertiary centre, underscoring the importance of contextualising quality improvement efforts according to baseline capacity and system readiness.

A key contribution of this work is demonstrating how AMS-IPC integration can be operationalised in LMIC hospital settings, corroborating previous literature on the value of coordinated governance and implementation. Rather than implementing separate and siloed programmes, integration was achieved through shared governance, coordinated action planning, and alignment of surveillance, training, and clinical feedback processes, particularly for the management of priority AMR pathogens [13,37]. Gains were most pronounced in domains driven by leadership engagement, coordination, and behavioural change, while areas requiring substantial financial investment, such as water safety and diagnostic capacity, showed limited short-term progress [7,15]. This highlights the need for differentiated expectations and timelines when evaluating integrated AMS-IPC interventions.

Compared with standalone AMS or IPC approaches, the integrated model enabled hospitals to address cross-cutting constraints affecting both programmes, including weak governance, insufficient training, and limited feedback to clinicians. These challenges have been widely reported across LMIC settings, where gaps in infrastructure, diagnostics, and workforce capacity hinder implementation [5,15,38]. Embedding stewardship within broader patient safety and infection control systems enhanced the relevance and uptake of AMS activities, while IPC practices aligned with AMS priorities benefited from stewardship input and leadership engagement, reinforcing the bidirectional relationship between antibiotic use and infection prevention [6].

The Fleming Fund investment played a critical enabling role by providing not only technical tools and training, but also a structured framework for reflection, prioritisation, and accountability. By leveraging existing surveillance and laboratory capacity, the project focused on strengthening implementation processes at the facility level. Consistent with prior evidence from Viet Nam and the wider Southeast Asian region, governance, leadership, and education emerged as key enablers of AMS implementation [14,26], while resource constraints, limited diagnostics, and workforce challenges remained persistent barriers [5,15]. Similar investments across Fleming Fund-supported countries have prioritised strengthening AMR surveillance, laboratory capacity, and data systems, while evidence of integrated AMS-IPC implementation at the facility level, particularly using structured quality improvement approaches, remains limited [39,40]. The network-based approach [25] facilitated cross-site learning and benchmarking, supporting more rapid uptake of effective practices than typically reported in single-site or short-term interventions.

Strengths of this study include its multisite design, use of validated tools, and explicit integration of AMS and IPC within a quality improvement framework. Limitations include the absence of a control group, short and variable follow-up periods, reliance on partly self-reported data, and inability to attribute observed improvements to downstream clinical or resistance outcomes within the project timeframe.

Overall, these findings suggest that integrated AMS-IPC strengthening, guided by repeated assessment and feedback, is feasible and scalable in hospitals with foundational structures in place. For national programmes and Fleming Fund-type investments, relatively modest, targeted support for implementation processes, alongside existing investments in surveillance and laboratory capacity, can yield substantial gains in AMS-IPC integration and support sustainable responses to antimicrobial resistance in LMICs. Our findings align with WHO guidance emphasising integration of AMS and IPC activities in resource-limited settings and extend previous work from Viet Nam and Southeast Asia demonstrating the feasibility of stewardship programmes despite persistent resource constraints [13,14,21].

In conclusion, this multisite quality improvement initiative demonstrates that integrating AMS and IPC through repeated assessment, structured feedback, and context-adapted action planning is feasible and associated with meaningful improvements in hospital AMS implementation across diverse settings in Viet Nam. By leveraging complementary assessment tools and a network-based support model, hospitals were able to translate surveillance capacity and policy guidance into coordinated, locally relevant actions. The findings highlight that impact of investments targeting governance, workforce capacity, and implementation processes, rather than infrastructure alone. This approach provides a scalable model for strengthening hospital responses to antimicrobial resistance in LMICs and offers important lessons for national programmes and Fleming Fund-type investments seeking sustainable impact.

## Data Availability

All data produced in the present study are available upon reasonable request to the authors.

## Patient and public involvement

Patients and the public were not involved in the design, conduct, reporting, or dissemination plans of this quality improvement initiative. The work focused on hospital-level systems and professional practice related to antimicrobial stewardship and infection prevention and control.

## Consent to participate

Not applicable. No individual patient data were collected, and participation involved healthcare professionals contributing to routine programme assessments and quality improvement activities.

## Data availability statement

Data generated and analysed during this study are available from the corresponding author on reasonable request. Data are not publicly available as they contain hospital-level programme information that could be sensitive if taken out of context.

## Author contributions

VTLH, TK, and HRvD contributed to the conception and design of the quality improvement initiative. Data collection and programme implementation were coordinated by the study team in collaboration with participating hospitals, supervised by VTLH, TK, NTC, TAC, NVM & VBD. NQP and NHY conducted data analysis. The manuscript was drafted by VTLH and critically reviewed and revised by all authors. All authors approved the final version of the manuscript.

## Acknowledgements

We thank the leadership, antimicrobial stewardship committees, infection prevention and control teams, microbiology laboratories, pharmacists, and clinical staff at the participating hospitals for their engagement and commitment to this quality improvement initiative. We also acknowledge the technical support provided by partners involved in the Fleming Fund programme in Viet Nam.

## Transparency statement

The authors affirm that this manuscript is an honest, accurate, and transparent account of the quality improvement work being reported; that no important aspects of the study have been omitted; and that any discrepancies from the original plan have been explained.

## Supplemental Data

**Table S1:**
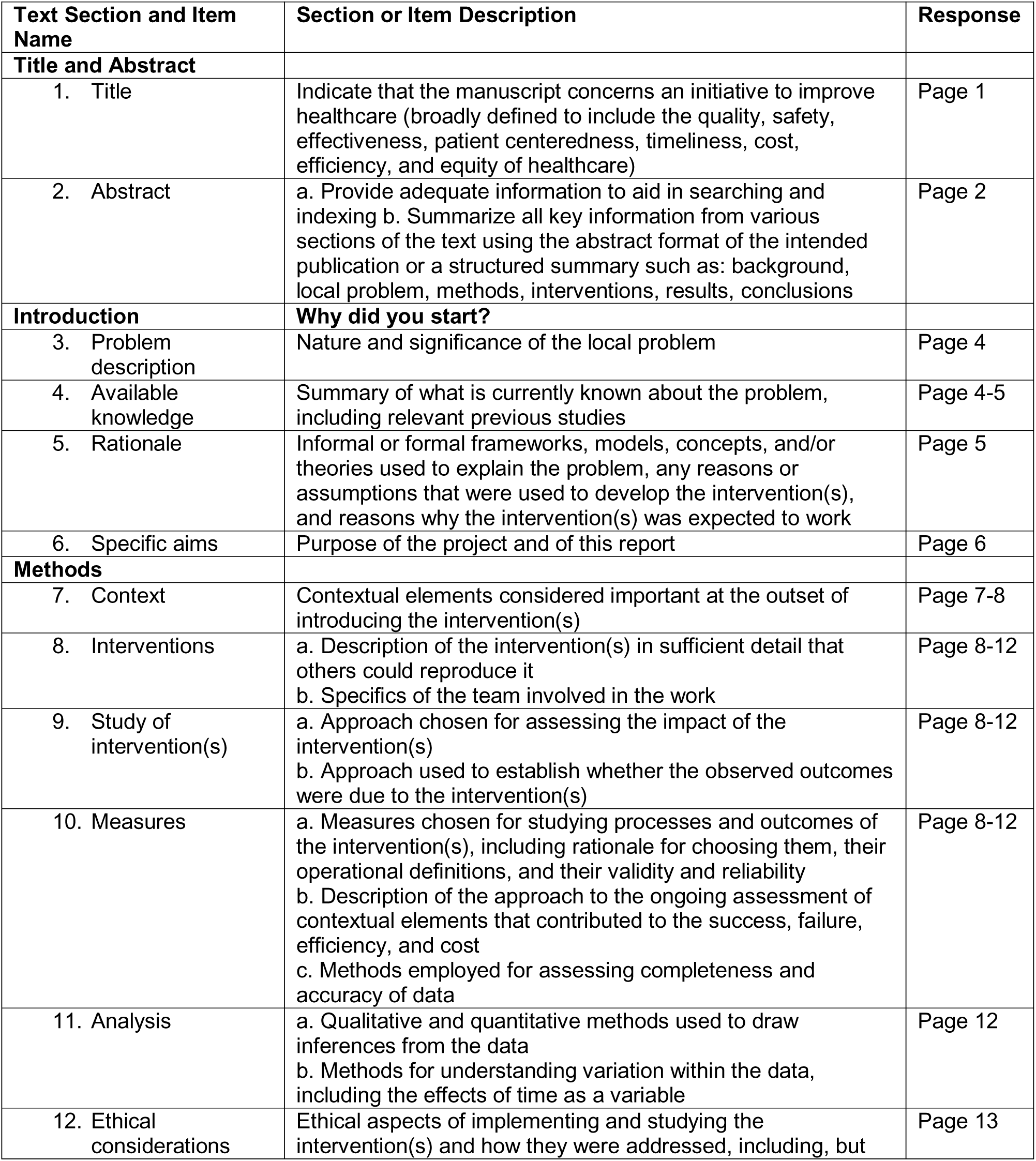
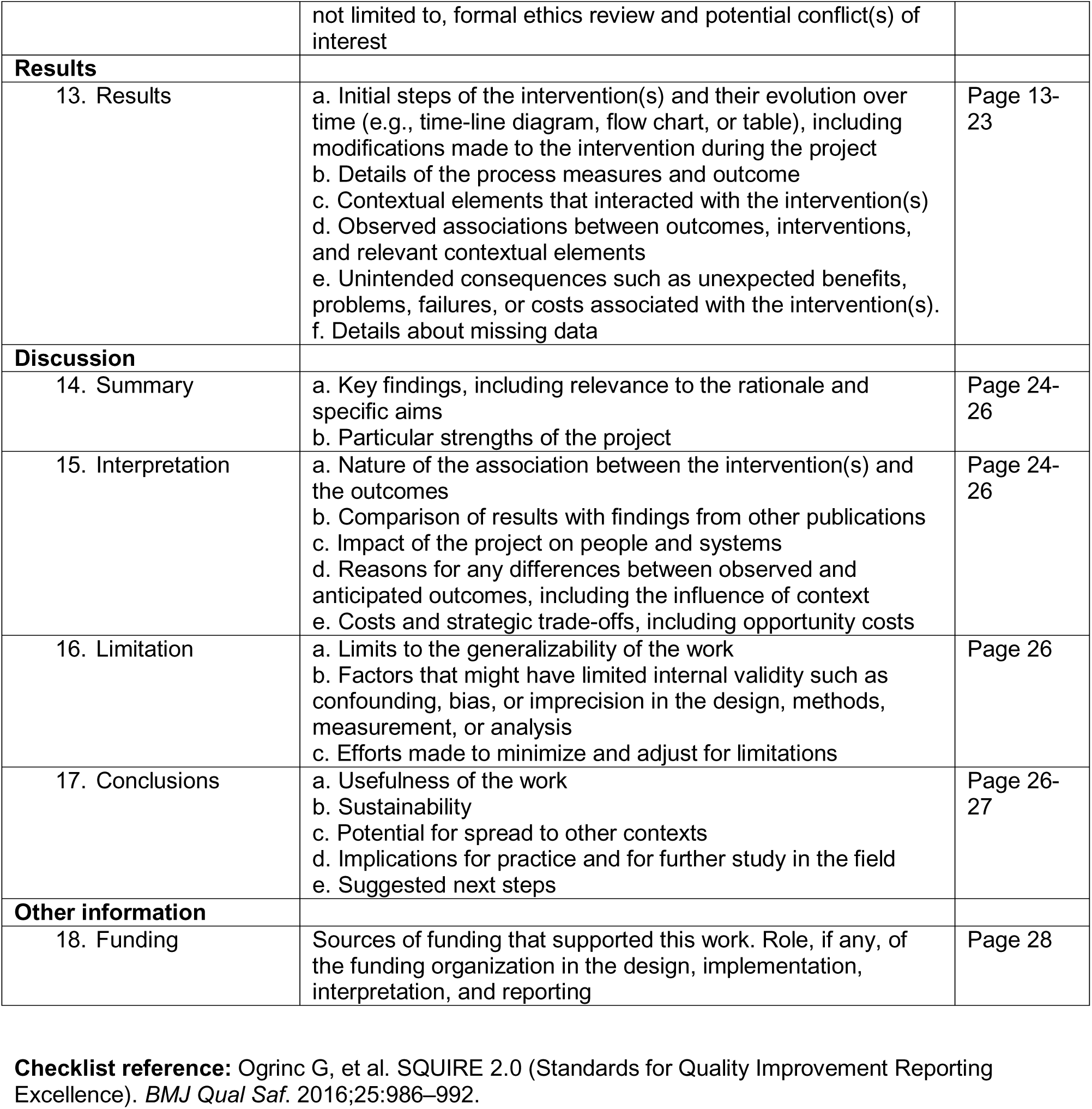
Revised Standards for Quality Improvement Reporting Excellence (SQUIRE 2.0)

## Supplemental Methods. Implementation of AMS in hospitals participating in the Fleming-funded project (2024–2025)

At each participating hospital, a structured process was implemented to support the development and execution of antimicrobial stewardship (AMS) programmes (Table S2). This process began with baseline AMS assessments and initiation meetings with hospital leadership to identify existing capacities and gaps. Action-planning meetings, conducted both face-to-face and online, were then used to co-develop hospital-specific AMS plans, followed by regular follow-up meetings to review progress and address implementation challenges. Draft action plans were developed and internally reviewed before being finalized and formally approved through hospital decision letters. Once approved, the plans were disseminated to relevant departments to support implementation. Key AMS activities, including training, audit and feedback, and treatment guideline updates, were subsequently initiated. Implementation progress was continuously monitored and supported by the Oxford University Clinical Research Unit (OUCRU) Viet Nam through ongoing engagement aligned with priorities identified during the planning process.

### Initiation and planning meetings with AMS committees

The research team conducted a series of structured engagement activities to assess and support AMS implementation at each site (Table S2). Initial meetings were held with hospital leadership and relevant stakeholders to review the status of AMS implementation and the functioning of AMS committees. From December 2024, two online meetings were conducted at each hospital: (1) a discussion with the coordination team and hospital leadership, and (2) a joint session with the coordination team and clinicians to introduce AMS activities and conduct a baseline AMS assessment. In addition, one in-person planning meeting was held at each hospital with either the hospital leadership and AMS committee or, where formal AMS structures were not yet established, with representatives from all departments.

AMS committees were encouraged to review ongoing AMS-related activities, including pharmacy reports, integrated plans, and infection prevention and control activities, and to conduct a self-assessed strengths, weaknesses, opportunities, and threats (SWOT) analysis for presentation and discussion during the meetings. Follow-up meetings were subsequently held to review progress and refine implementation plans. Hospitals were also advised to update AMS committee membership, clarify roles and responsibilities, and develop a one-year action plan (to be annually reviewed and updated). Expected outputs included hospital-specific AMS action plans for 2025–2026, focusing on key activities such as staff training, audit and feedback, standardized reporting, and updating treatment guidelines in line with revised national AMS guidance.

### AMS workshops

Based on the outcomes of these discussions, OUCRU Viet Nam delivered four targeted workshops to address locally identified needs. Workshop content was informed by the priorities identified by AMS committees and teams, as well as findings from the knowledge, attitudes, and practices (KAP) survey. All workshops were delivered online. Participation rates were monitored, and engagement was assessed by the proportion of designated staff attending each workshop.

The poster of AMS online workshops*:

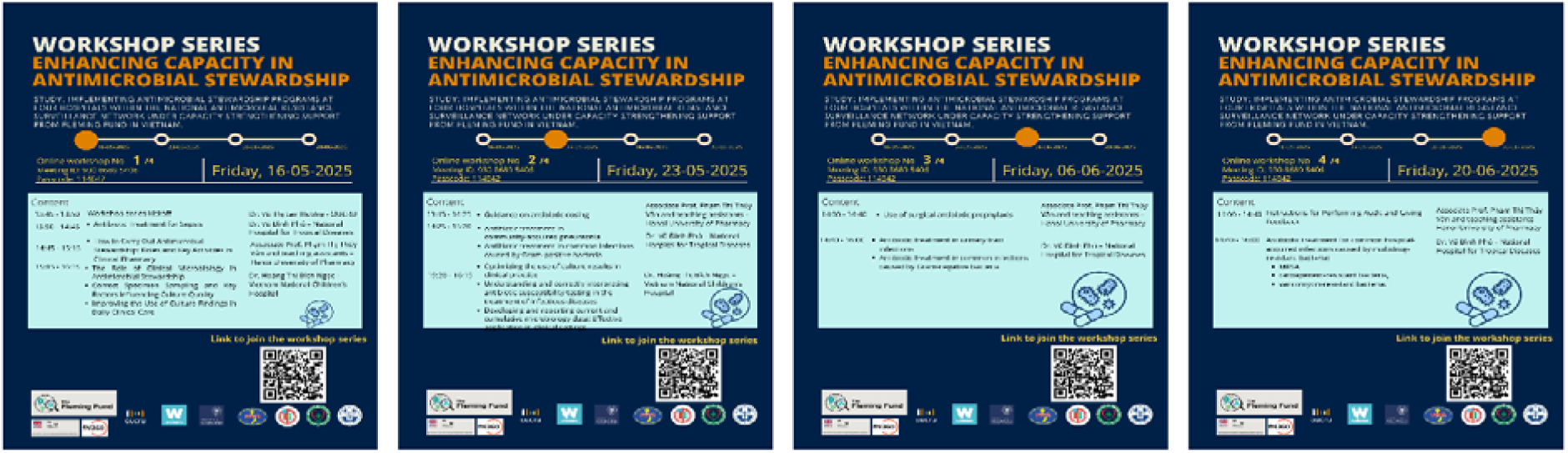

### Supportive documents and standard operating procedures

Online resources outlining standard operating procedures (SOPs) for AMS activities were provided to all participating hospitals. These SOPs were updated in line with national policy, including Decision No. 5631/QD-BYT, and relevant international guidance, and were used as reference materials to support local implementation.

SOPs included:

- audit and feedback procedures linked to the antibiotic prescribing systems
- intravenous-to-oral switch (IV-to-PO) procedures
- lists of authorized antibiotics and approval processes
- AWaRe guidance
- AMS leaflets and posters
- an antibiogram template and development procedures
- an AMS reporting template for the four participating hospitals

AMS information, education, and communication (IEC) materials included:

- IV-to-PO posters
- materials on authorized antibiotic lists and approval processes
- AWaRe guideline posters and AWaRe classification lists
- educational leaflets and posters for doctors, patients, and caregivers

### Treatment guidelines

Hospital AMS committees, in collaboration with OUCRU Viet Nam and the National Hospital for Tropical Diseases (NHTD), reviewed existing treatment guidelines and updated them where necessary to align with current best practices and local evidence. The revised guidelines were disseminated to participating hospitals. Hospitals were also strongly encouraged to proactively review and update their local treatment guidelines to support audit and feedback activities for antibiotic prescribing. All participating hospitals undertook such reviews and included the updating of key treatment guidelines in their annual action plans, including guidelines related to surgical antibiotic prophylaxis.

**Table S2.**
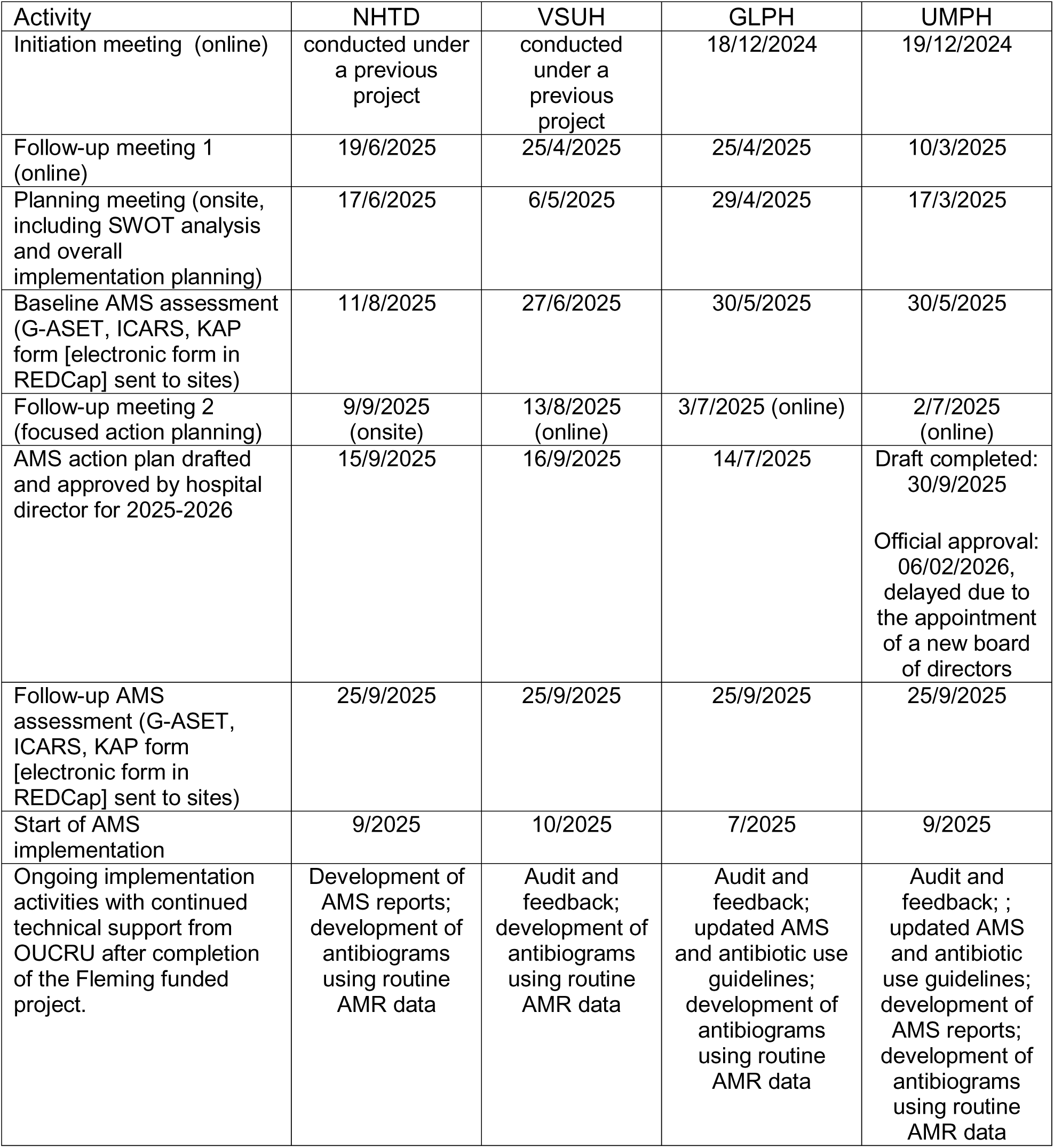
Timeline of antimicrobial stewardship (AMS) activities across participating hospitals, 2024–2025.

| Activity | NHTD | VSUH | GLPH | UMPH |
| --- | --- | --- | --- | --- |
| Initiation meeting (online) | conducted under a previous project | conducted under a previous project | 18/12/2024 | 19/12/2024 |
| Follow-up meeting 1 (online) | 19/6/2025 | 25/4/2025 | 25/4/2025 | 10/3/2025 |
| Planning meeting (onsite, including SWOT analysis and overall implementation planning) | 17/6/2025 | 6/5/2025 | 29/4/2025 | 17/3/2025 |
| Baseline AMS assessment (G-ASET, ICARS, KAP form [electronic form in REDCap] sent to sites) | 11/8/2025 | 27/6/2025 | 30/5/2025 | 30/5/2025 |
| Follow-up meeting 2 (focused action planning) | 9/9/2025 (onsite) | 13/8/2025 (online) | 3/7/2025 (online) | 2/7/2025 (online) |
| AMS action plan drafted and approved by hospital director for 2025-2026 | 15/9/2025 | 16/9/2025 | 14/7/2025 | Draft completed: 30/9/2025<br>Official approval: 06/02/2026, delayed due to the appointment of a new board of directors |
| Follow-up AMS assessment (G-ASET, ICARS, KAP form [electronic form in REDCap] sent to sites) | 25/9/2025 | 25/9/2025 | 25/9/2025 | 25/9/2025 |
| Start of AMS implementation | 9/2025 | 10/2025 | 7/2025 | 9/2025 |
| Ongoing implementation activities with continued technical support from OUCRU after completion of the Fleming funded project. | Development of AMS reports; development of antibiograms using routine AMR data | Audit and feedback; development of antibiograms using routine AMR data | Audit and feedback; updated AMS and antibiotic use guidelines; development of antibiograms using routine AMR data | Audit and feedback; ; updated AMS and antibiotic use guidelines; development of AMS reports; development of antibiograms using routine AMR data |

**Figure S1.**
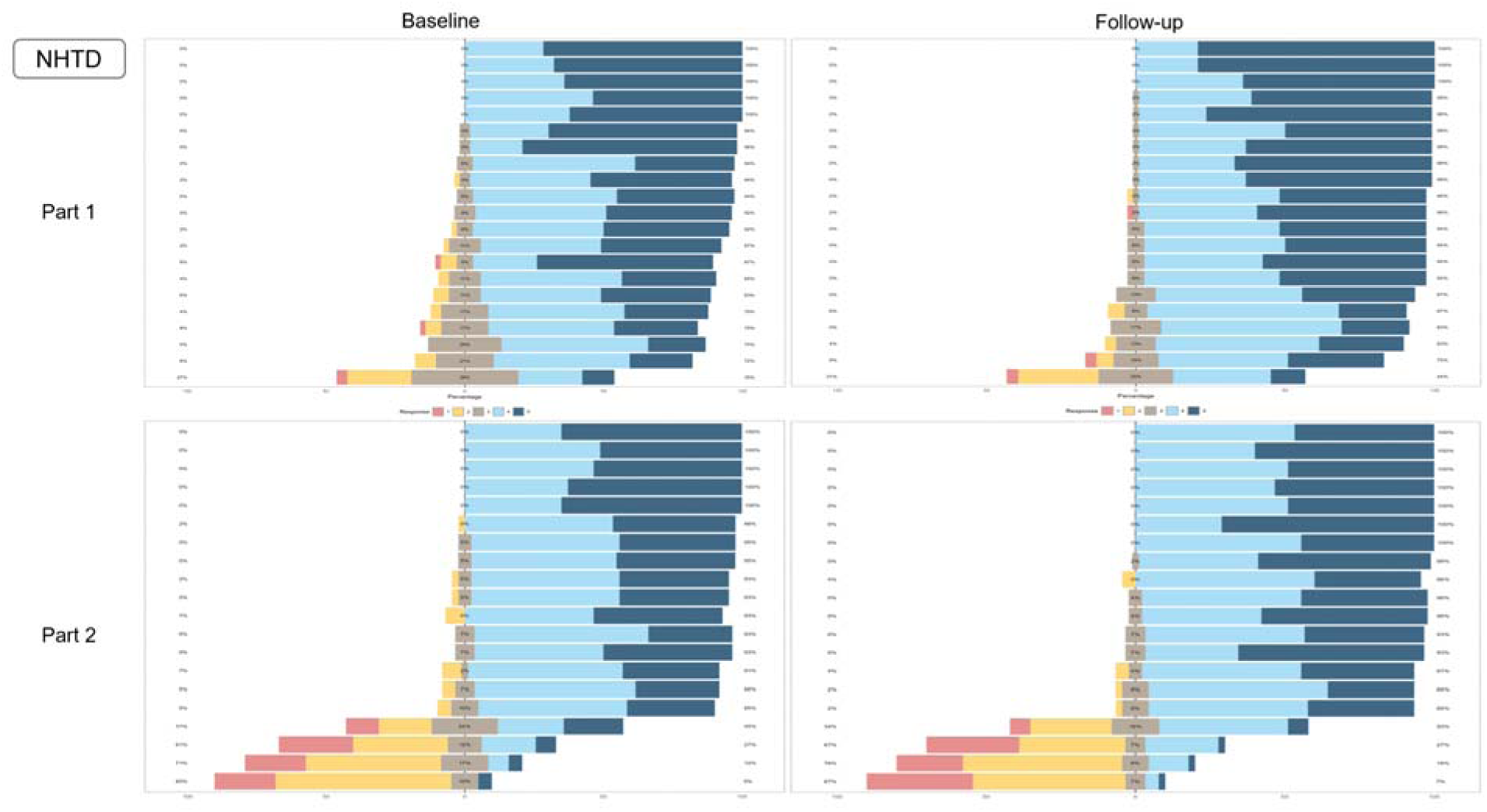
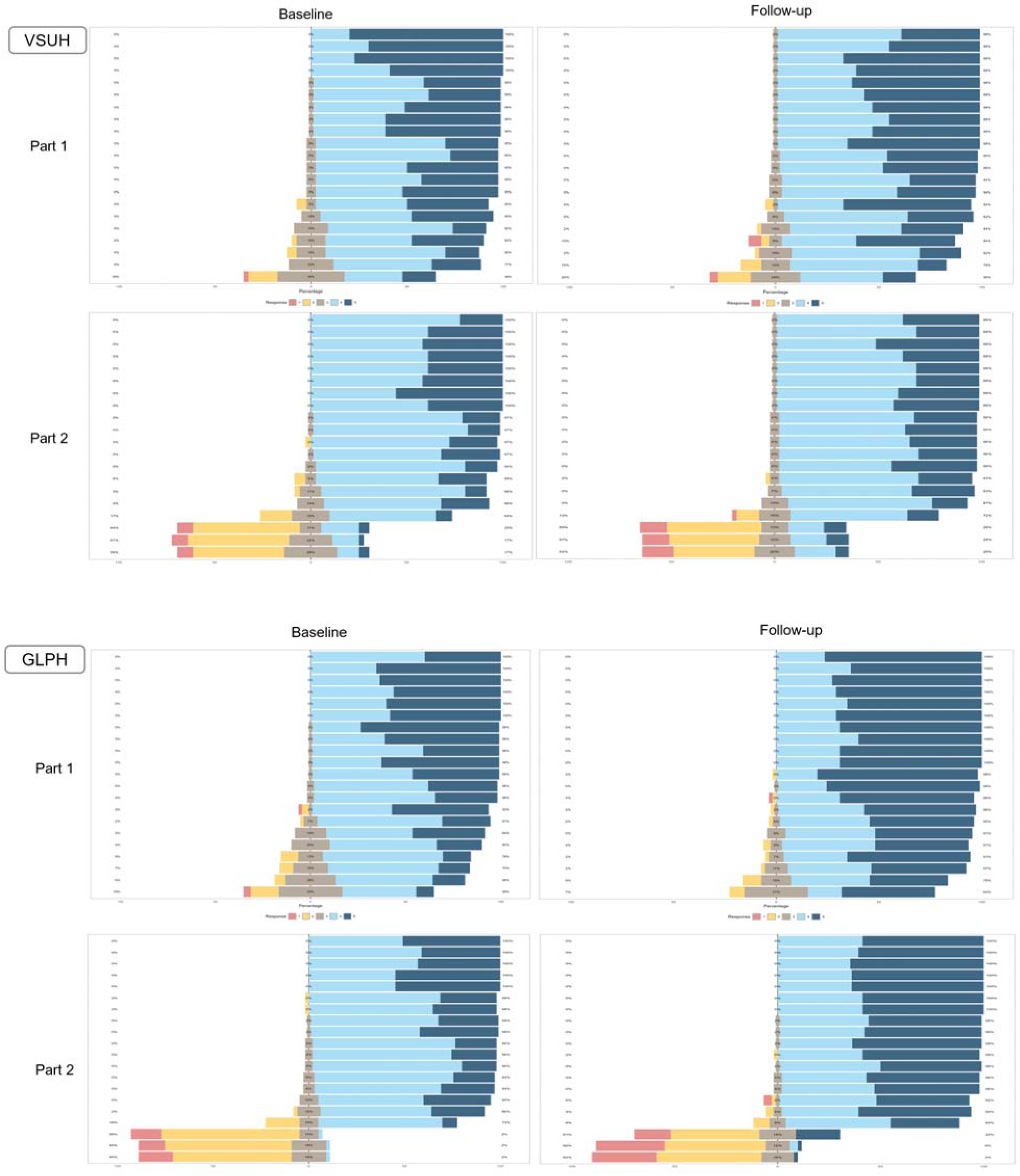
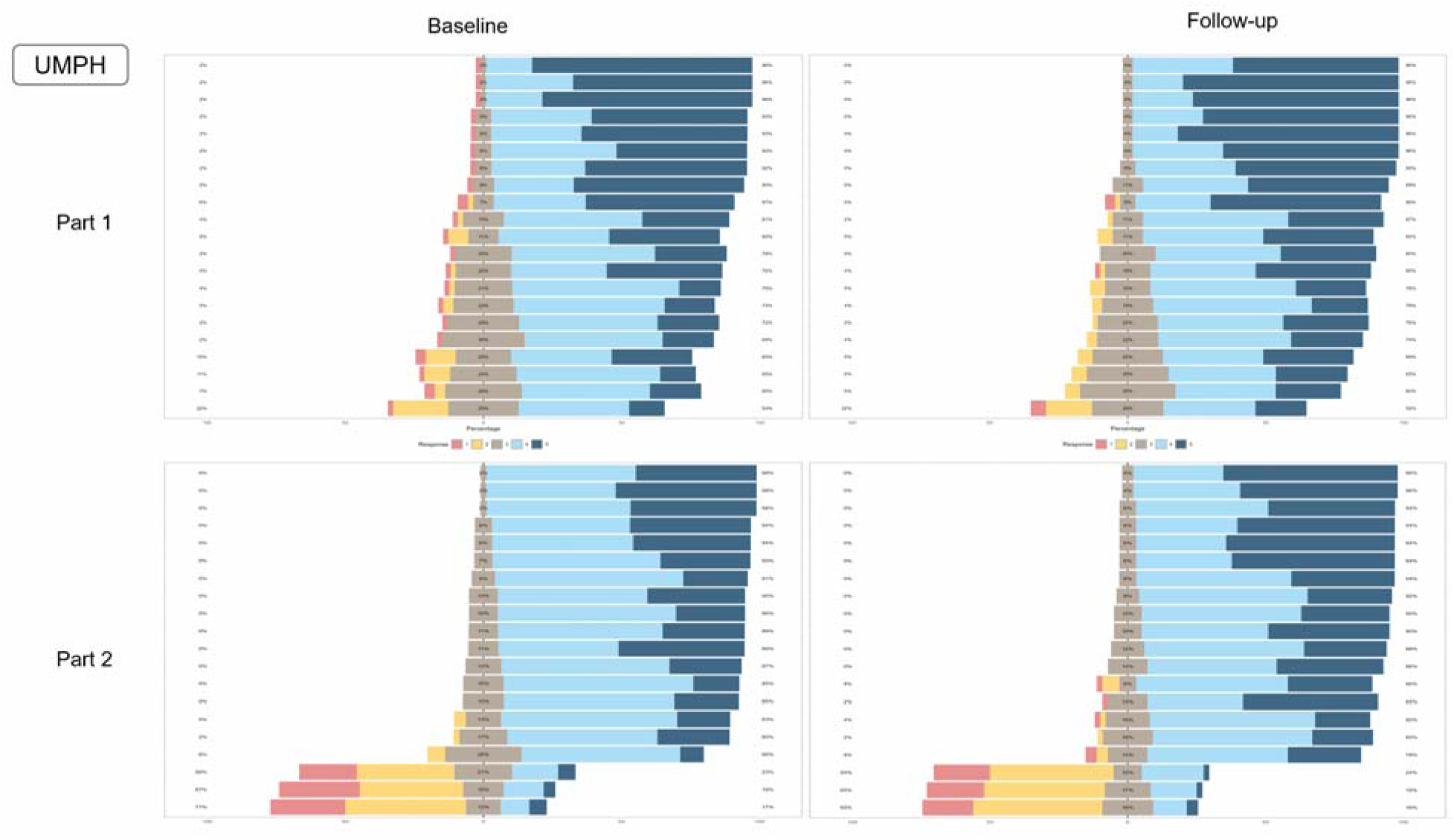
Visualisations of the KAP results were used to support discussion of targeted training priorities during AMS team meetings. At each timepoint, each hospital administered the KAP survey to approximately 50 staff members, including doctors, pharmacists, and microbiologists, with participation determined by staff availability at the time of assessment. The survey was based on the KAP form developed within the US CDC’s G-ASET toolkit [32,33]. The figures suggested small but consistent shifts towards more favourable responses at follow-up, particularly for attitude and practice items, although the magnitude and pattern of change varied across hospitals and several items remained unchanged. These observations were based on visual inspection rather than formal statistical inference and should therefore be interpreted cautiously. Nonetheless, the KAP surveys were useful in stimulating discussion and informing education and training activities to support programme implementation.

